# Comparing mental health trajectories of four different types of key workers with non-key workers: A 12-month follow-up observational study of 21,874 adults in England during the COVID-19 pandemic

**DOI:** 10.1101/2021.04.20.21255817

**Authors:** Elise Paul, Hei Wan Mak, Daisy Fancourt, Feifei Bu

## Abstract

**Background:** There are concerns that key workers may be at a greater risk for psychological distress than non-key workers during the COVID-19 pandemic. However, little research has included key workers outside of the healthcare sector or has disaggregated key workers into different subgroups.

**Aims:** To examine longitudinal changes in mental health over 12 months during the COVID-19 pandemic comparing four different groups of key workers with non-key workers.

**Method:** Longitudinal data were from 21,874 adults living in England (21 March 2020 to 22 February 2021). Latent growth modelling (LGM) was utilised to compare growth trajectories of depressive and anxiety symptoms in non-key workers and four types of key workers: i) health and social care workers, ii) teachers and childcare workers, iii) public service workers, and iv) essential services key workers (e.g., food chain or utility workers).

**Results:** When accounting for both time-invariant and time-varying covariates, key workers in the essential services category had consistently higher levels of depressive and anxiety symptoms than non-key workers across the whole of the study period. There was little difference in mental health trajectories between health/social care, teachers/childcare and public service worker categories and non-key workers.

**Conclusions:** Our findings suggest risk for poorer mental health during the COVID-19 pandemic varies within the broad category of key workers generally, and that those working in utility, food chain, and transport roles are especially at risk. Future research should focus on identifying which aspects of working conditions may be contributing to occupational stress in these groups.

## Introduction

The COVID-19 pandemic has had substantial detrimental effects on public mental health.^1^ Certain subgroups such as key workers (e.g., individuals working in the health care and social support sectors, delivery workers, teachers) have been posited to be more adversely affected than the rest of the population.^2^ However, despite key workers comprising a significant proportion of the population (33% in the UK),^3^ and fulfilling a large variety of roles with differing levels of exposure to the public and thus to the virus itself,^4^ the majority of the research on key worker mental health has focused exclusively on healthcare workers^5,6^ or has examined key workers broadly as a collective.^7–10^ Key workers in general have been found to be more likely than non-key workers to meet criteria for clinically significant mental distress^9^ and probable criteria for depression, anxiety, and post-traumatic stress disorders than non-key workers.^11^ However, other studies conducted during the COVID-19 pandemic have not found key workers to report more depressive or anxiety symptoms than non-key workers.^7,8,12^ These equivocal findings may be due to the heterogeneity involved when grouping all key workers together. For example, even amongst the specific category of healthcare workers, those whose jobs require direct contact with COVID-19 patients display more symptoms of anxiety, depression, insomnia and traumatic stress than healthcare workers not working directly with COVID-19 patients.^5,6^

These varied findings may also be due to variation in the amount of stress experienced across other key worker groups in different sectors. The mental health of key workers fulfilling roles other than in healthcare may have been disproportionately impacted during the current pandemic for several reasons. First, like healthcare workers, they have also been required to leave their homes for work despite the associated risk of infection and mortality in themselves and in family and friends.^4,13^ They may have also been challenged by longer working hours and more intense working circumstances, sometimes with inadequate personal protective equipment (PPE),^14,15^ the latter of which has been found to be associated with depressive and anxiety symptoms amongst key workers during the current pandemic.^16^ Degree of exposure to the public and therefore to the virus may also correlate with increased distress in other key worker roles as well. One US study conducted in the first few months of the pandemic found that grocery store workers who interacted directly with the public were more anxiety and depressed than those who do not interface with customers.^17^ Some key workers may be additionally have already been at greater risk for severe infection and death from COVID-19 due to age, pre-existing health conditions, living in areas of high socioeconomic deprivation, and belonging to an ethnic minority group.^4,13^

Only two studies have disaggregated and compared the mental health of different key worker groups other than in the healthcare sector. In the first few months of the pandemic in the UK, one study found that food workers were the only key worker group of the eight groups studied more likely than the others (e.g., utility workers, transport workers, health and social care workers) to meet probable criteria for anxiety disorder.^11^ In the same study, all key worker groups besides transport workers were more likely to meet criteria for probable post-traumatic stress disorder (PTSD) than non-key workers, and odds for meeting this criterion varied substantially, from 1.7 in health and social care workers to 3.4 in public service workers.^11^ A second study conducted in Australia reported key workers other than in the healthcare sector to have higher levels of anxiety, depression, stress, and poorer quality of life than healthcare workers and the rest of the population.^18^

Together, these findings suggest that while key workers broadly may be more vulnerable to experiencing poorer mental health than the rest of the population during the current pandemic,^9,11^ key workers in other roles may be more at risk than those in the health care sector.^11,18^ However, research on the longitudinal changes of mental health of the key workers as the COVID-19 pandemic develops is lacking. It is also unknown whether mental health trajectories vary amongst different key worker groups. Therefore, the aim of this study was to compare the growth trajectories of anxiety and depressive symptoms of four categories of key workers with non-key workers over the first 12 months (March 2020 to February 2021) of the pandemic in the UK. The findings will inform our understanding of which specific key worker groups may be most in need of psychosocial support during the current and in future pandemics. Further, given that the effects of the COVID-19 pandemic on mental health could be long-lasting, it is crucial to identify various mental health experiences amongst key workers that could help in designing policies and implementations to support those who may be continued to be affected by the COVID-19 crisis in post-pandemic times.

## Methods

### Study design and participants

This study analysed data from the University College London (UCL) COVID-19 Social Study; a large panel study of the psychological and social experiences of over 75,000 adults (aged 18+) in the UK during the COVID-19 pandemic. The study commenced on 21 March 2020 and involves weekly and then monthly (four-weekly) online data collection from participants for the duration of the pandemic. The study did not use a random sample design and therefore the original sample is not representative of the UK population. However, it does contain a heterogeneous sample that was recruited using three primary approaches. First, convenience sampling was used, including promoting the study through existing networks and mailing lists (including large databases of adults who had previously consented to be involved in health research across the UK), print and digital media coverage, and social media. Second, more targeted recruitment was undertaken focusing on (i) individuals from a low-income background, (ii) individuals with no or few educational qualifications, and (iii) individuals who were unemployed. Third, the study was promoted via partnerships with third sector organisations to vulnerable groups, including adults with pre-existing mental health conditions, older adults, carers, and people experiencing domestic violence or abuse. The study was approved by the UCL Research Ethics Committee [12467/005] and all participants gave informed consent. A full protocol for the study is available online at www.COVIDSocialStudy.org.

In the present study, we restricted the sample to participants who were in employment at baseline and living in England (N= 39,255). Further, we included only participants with at least three repeated measures between 21 March 2020 and 22 February 2021. These criteria provided us with data from 22,012 participants. Around 1% of these participants withheld data or preferred not to self-identify on demographic and health-related factors and were therefore excluded from our analysis. This provided us with a final analytic sample size of 21,874 participants who were followed up for a maximum of 12 months.

### Measures

#### Key worker status

When participants first joined the study, they were asked if they were currently fulfilling any of the government’s nine identified key worker roles. These were categorised into five groups: (1) non-key worker (reference category), (2) health, social care or relevant related support worker, (3) teacher or childcare worker, (4) public service worker (e.g., justice staff, religious staff, public service journalist or mortuary worker, local or national government worker), and (5) essential services key worker (e.g., food chain, utility, public safety or national security worker, worker involved in medicines or protective equipment production or distribution).

#### Outcome variables

Depressive symptoms were measured using the Patient Health Questionnaire (PHQ-9);^19^ a standardised instrument for screening for depression in primary care. Unlike the original PHQ-9, the current study enquired about symptoms ‘over the last week’ instead of ‘over the last two weeks’ as data were initially collected weekly. The questionnaire includes 9 items with 4-point responses ranging from ‘not at all’ to ‘nearly every day’. Higher overall scores indicate more depressive symptoms, ranging from 0 to 27. Anxiety symptoms were measured using the Generalized Anxiety Disorder assessment (GAD-7);^20^ a well-validated tool used to screen for Generalized Anxiety Disorder in clinical practice and research. These questions were also worded as ‘over the last week’ for the same reason as the depression items. The GAD-7 comprises 7 items with 4-point responses ranging from ‘not at all’ to ‘nearly every day’, with higher overall scores indicating more symptoms of anxiety, ranging from 0 to 21. These data were collected weekly between 21 March and 21 August 2020, and then monthly (four-weekly) starting from 24 August 2020 onwards (see Table S1). Our analyses used months as the unit of time. Mean values of depressive and anxiety symptoms across four weeks were used when data were collected weekly.

#### Time-invariant covariates

A range of socio-demographic and health factors were considered as potential confounders. These included gender (women vs men), ethnicity (white vs ethnic minorities), age groups (age 18-29, 30-45, 46-59, 60+) and education (up to GCSE levels, A-levels or equivalent, and university degree or above). We included two health-related factors: self-reported diagnosis of any long-term physical health condition (e.g., asthma or diabetes) or any disability (yes vs no), and self-reported diagnosis of any long-term mental health condition (e.g., depression, anxiety) (yes vs no). All of these were time-invariant covariates that were measured when participants first joined the study.

#### Time-varying covariates

First, we considered a time-varying covariate to indicate if participants had gone to work outside of the home (yes/no). To provide consistent measurement between the weeks of the study when participation was weekly vs monthly, we coded this variable as whether participants worked outside at any point during the prior four-week period. Second, in December 2020 the UK began its COVID-19 vaccination programme and health and social care workers were one of the priority groups to be vaccinated. Therefore, we considered another time-varying covariate indicating if participants had had their first dose of the vaccine (yes/no) from December 2020 onwards (months 10-12).

#### Statistical analysis

Data were analysed using the latent growth modelling (LGM) approach. We used unspecified LGM which allows the shape of growth trajectories to be determined by data by using free time scores. In this model, two time scores were fixed at 0 and 1 respectively for the purpose of model identification, while the rest were estimated freely, allowing for an empirically based nonlinear shape for the outcome growth trajectory. In models for depressive and anxiety symptoms respectively, key worker status and other time-invariant covariates were allowed to predict the growth factors (intercept and slope; Model I). Then, we added the time-varying covariates allowing them to predict the outcomes directly (Model II). The full model specification is presented in Figure S1 in the Supplementary Materials.

Weights were applied throughout the analyses. The final analytical sample was weighted to the proportions of gender, age, ethnicity and education among adults in employment in England based on the Quarterly Labour Force Survey (March-May 2020).^21^ Main analyses were implemented in Mplus Version 8.

## Results

### Descriptive characteristics

In the unweighted analytic sample of 21,874 participants, women (78.8%) and people with a universiy degree or above (75.2%) were overrepresented, whereas younger adults (aged 18-29; 6.7%) and people from ethnic minority groups (5.4%) were underrepresented (Table 1). After weighting, the sample reflected population proportions, with 51.0% women, 39.4% participants with a university degree or above, 15.3% aged under 30, and 10.9% participants belonging to an ethnic minority group. Noticably, demographic and health charateristics differed across the key worker groups. For example, while there were more women than men in the ‘teacher/childcare’ category (74.2% vs 25.8%) and in the ‘health/social care’ category (69.8% vs 30.2%), the gender proportions in non-key worker and public service workers were similar. Only 15.0% of key workers in the essential services category had a degree or above, which was 35.8% amongst public service key workers, 41.9% amongst non-key workers, 47.1% amongst health and social care workers and 51.6% amongest teachers or childcare workers.

**Table 1.**
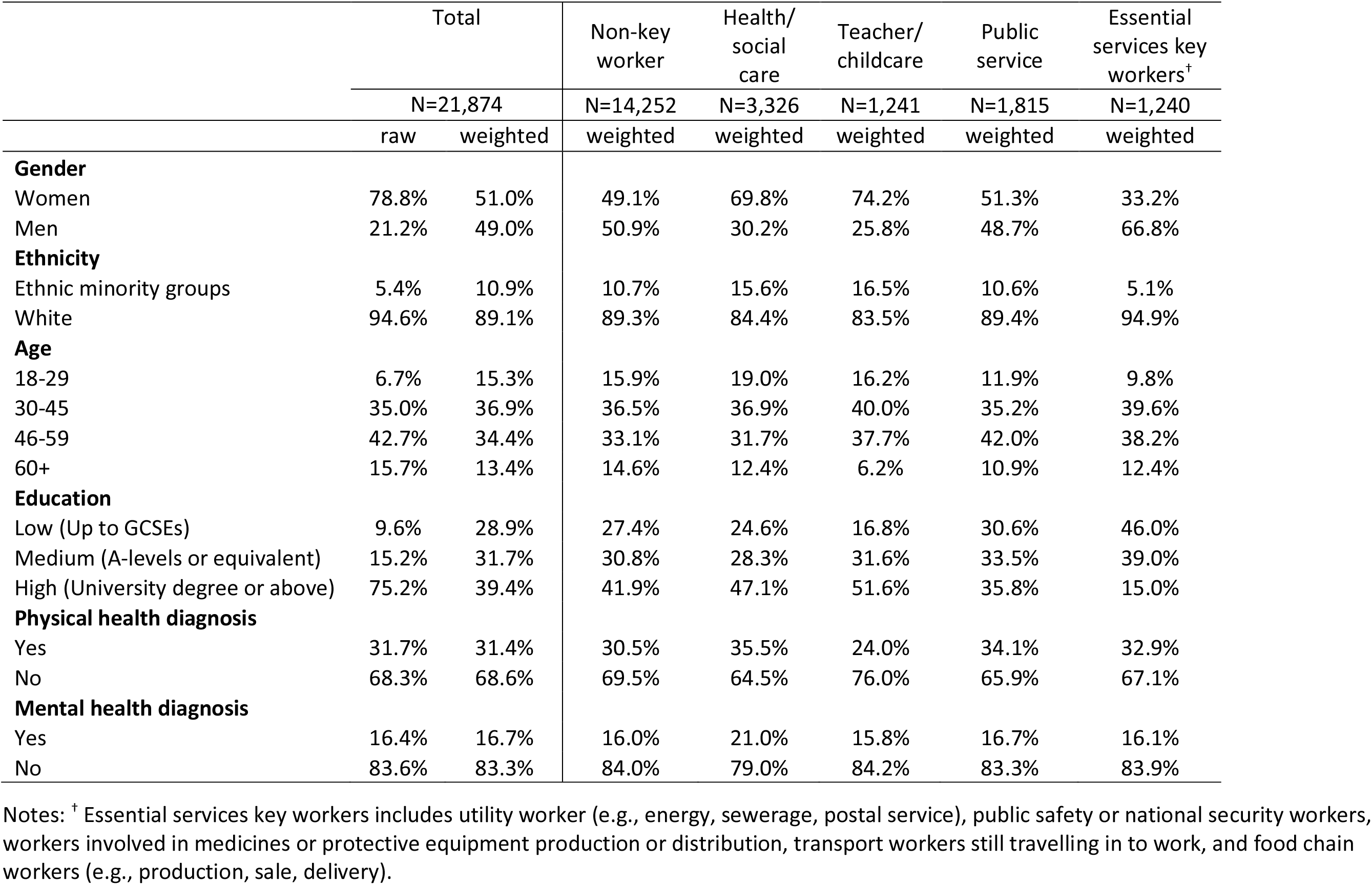
Descriptive statistics of the total analytical sample and by key worker status (weighted)

Figures 1a and 1b depict changes in the mean level of depressive and anxiety symptoms respectively and suggest some differences in the longitudinal changes in mental health by key worker status. Figure 1c shows the pecentages of participants who went outside their homes for work in each key worker category across different time points. Descriptive statistics of time-varying variables are shown in Figure 1. Generally speaking, key workers were more likely to have left home for work than non-key workers, especially teachers (when schools reopened in the Autumn 2020) and key workers in the essential services category. The latter group seemed to have left home for work most consistently over the course of the study period. As expected, the percentage of those who were vaccinated from December 2020 onwards increased dramatically for health and social care workers (Figure 1d). By contast, key workers in the essential services category had the lowest level of vaccination across all groups including non-key workers, with the gaps apprearing to widen over time.

**Figure 1.**
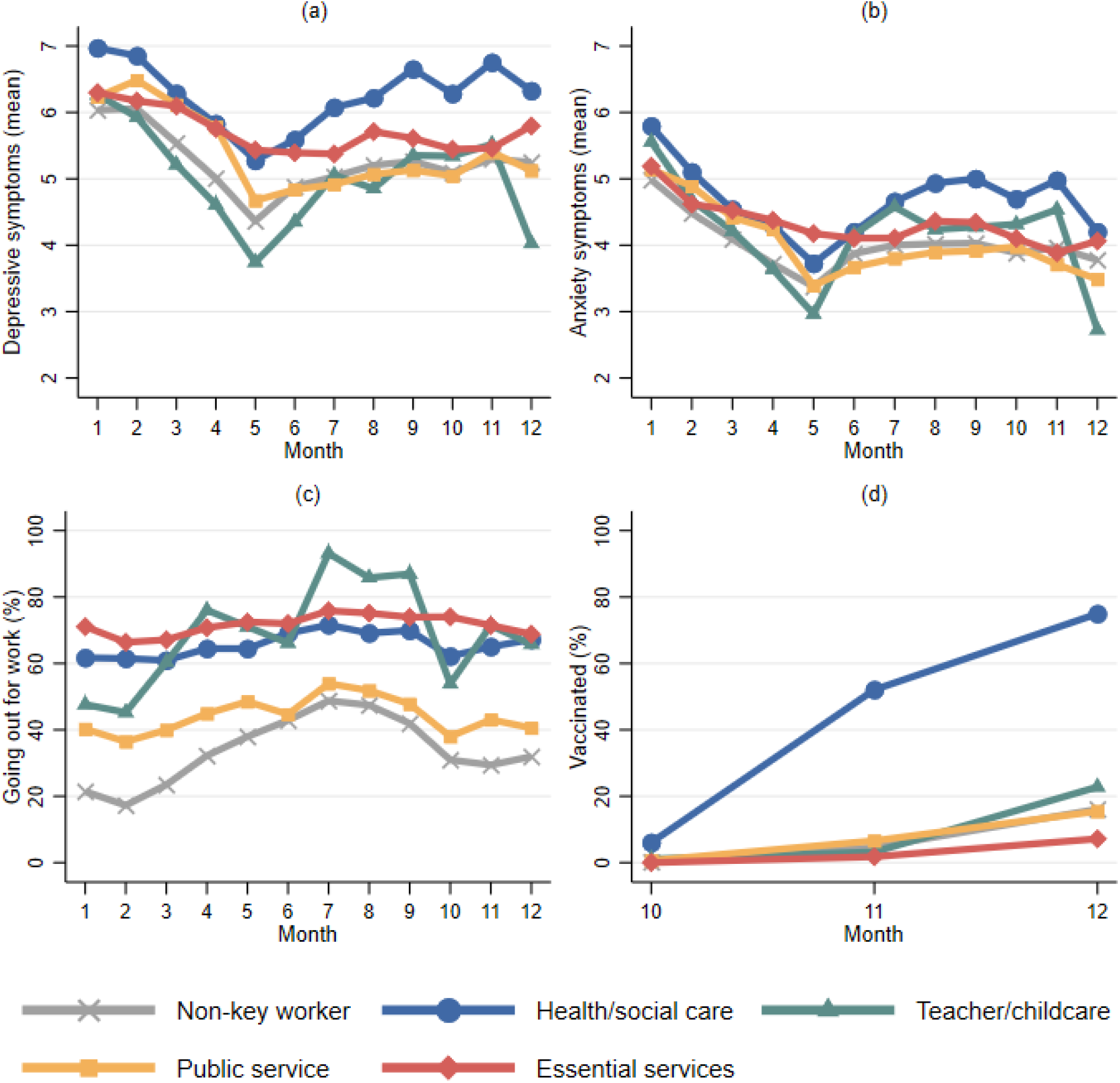
Descriptive statistics of the time-varying variables across time points by key worker status.

### Latent growth modelling

Overall, depressive and anxiety symptoms were worst at the start of the pandemic but then improved as restrictions from the first lockdown eased over the summer before worsening as cases increased again in the autumn of 2020. There was little evidence that the intercept or slope of depressive symptom growth trajectories differed by key worker status when only controlling for time-invariant socio-demographic and health factors (Figure 2a, see also Table S2 in the Supplementary Materials for full results). However, after controlling for the time-varying factors (leaving the house for work and COVID-19 vaccination status), key workers in the essential services category had significantly higher depressive symptoms at the start of the observational period (intercept) compared with non-key workers (Figure 2b). This was mainly due to the introduction of leaving the home for work, which was associated with higher levels of depressive symptoms at the start (month one) but reduced depressive symptoms between months four and seven (Table S2). There was no evidence that having had the COVID-19 vaccine was associated with changes in depressive symptoms. The difference in the intercept between teacher/childcare workers and non-key workers was also statistically significant. Specifically, teacher/childcare workers had fewer depressive symptoms at the start of the study. There was little difference in slopes across key worker status. Key workers in the essential services category had more, and teacher/childcare workers had fewer depressive symptoms than non-key workers consistently across the 12-month observational period.

**Figure 2.**
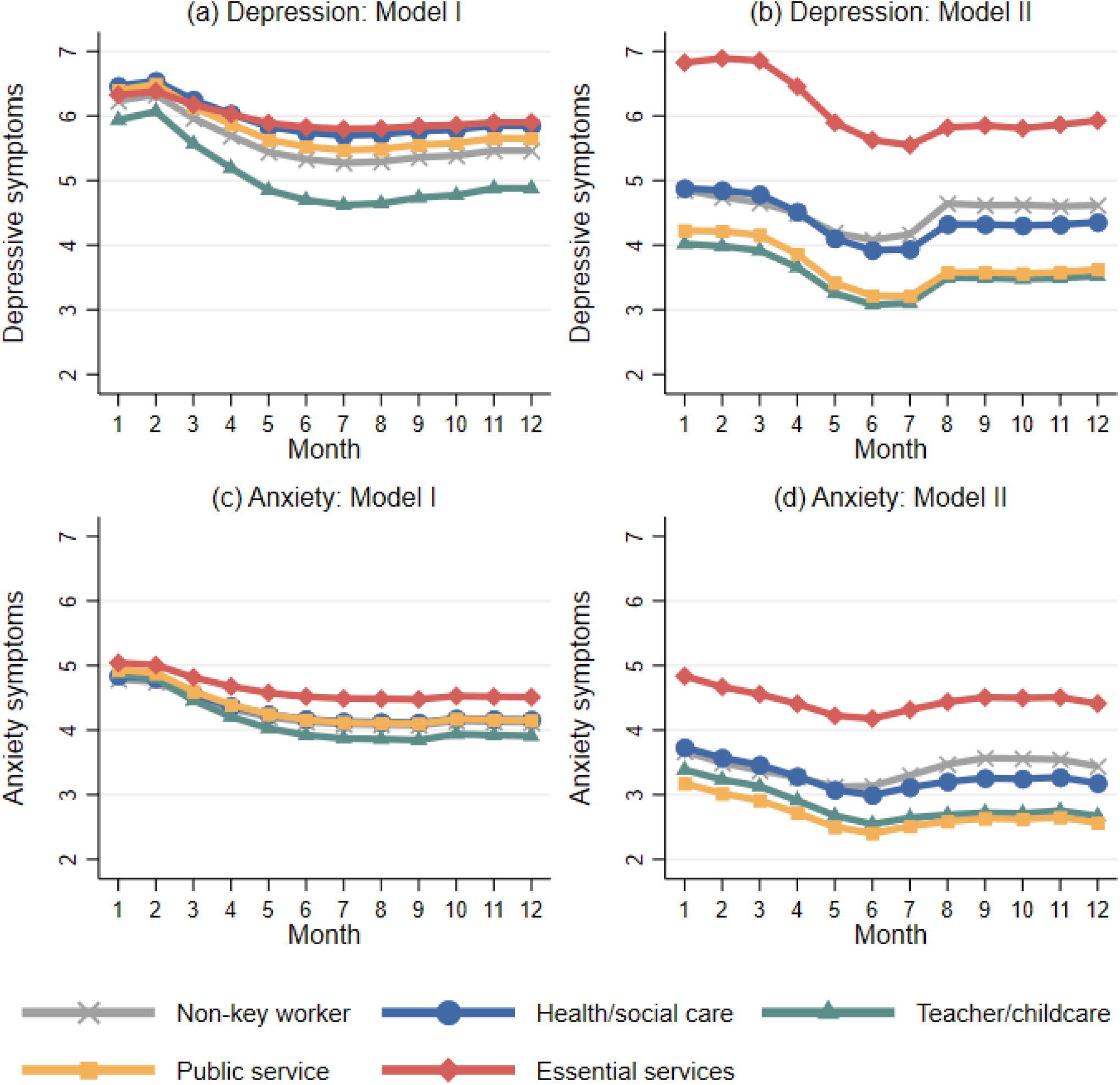
Predicted growth trajectories of depressive and anxiety symptoms by key worker status from latent growth models. Notes: Model I controlled for time-invariant covariates; Model II controlled for both time-invariant and varying covariates

The growth trajectories for anxiety symptoms were similar between non-key workers and each of the key worker categories when only including time-invariant covariates (Figure 2c). However, after controlling for time-varying covariates, in particular leaving the house for work, key workers in the essential services category had more anxiety symptoms compared with non-key workers and the other key worker groups across the entire study period. There was little difference in the slopes across the remaining key worker groups. Similar to the results for depressive symptoms, essential services key workers had a consistently higher score of anxiety symptoms than non-key workers across the whole of the observation period.

## Discussion

Our results show that key workers in essential service sectors (e.g., food chain, utility, transport, and public security or safety) had consistently higher depressive and anxiety symptoms over the entire one-year study period. In contrast, teacher/childcare workers had fewer depressive symptoms than non-key workers at the start and for the duration of the study. Despite the disproportionate research attention paid to the mental health of healthcare key workers during the pandemic, we did not find evidence that those working in the ‘health/social care’ and ‘public service’ sectors had levels of depressive and anxiety symptoms that were higher than the general population.

This is now the third study to have found worse mental health outcomes in key workers employed in sectors other than healthcare.^11,18^ In our study, key workers within essential services reported consistently, on average, 2-point higher levels of depressive symptoms and 1-point higher anxiety symptoms than non-key workers independent of potential confounding factors. There are a number of potential explanations for why this group may have been particularly badly affected. First, qualitative studies of such key workers have documented particular challenges such as adapting to duties under more stressful circumstances (e.g., increased workloads and fears of transmitting the virus to family members^15^). Second, individuals in essential services key worker roles (e.g., transport, postal, and retail workers) have received less recognition from the public or their employers for their efforts, which could have resulted in a feeling of inadequacy and further exacerbate their mental health.^15^ Third, essential services key workers are disproportionally more likely to have lower levels of educational attainment, to be in more routine occupational roles, and to experience financial hardship than other categories of key workers and the general population. Studies focusing on the experiences of people in roles of lower socio-economic position have consistently shown poor mental as well as physical health,^22^ these findings could therefore reflect existing socio-economic health inequalities within society.^23^ Finally, key workers in essential services roles have also been at particularly high risk for contracting COVID-19.^4,13^ It is notable that the distinctions between the mental health experiences of essential services key workers and other key worker roles were exacerbated when taking into account having to work outside the home, suggesting that exposure to risk may have had an adverse effect on mental health. This theory is also supported by qualitative work highlighting the challenges faced by essential services key workers in often being unable to access personal protective equipment (PPE) through their employers in comparison to other key worker groups such as health/social care workers and teachers where although challenges with PPE remained, there was more attention given to safe working environments.^15^

Another key finding from this study was that health and social care workers did not show higher levels of anxiety or depressive symptoms than non-key workers, although their levels were descriptively higher than some other groups such as teachers/childcare key workers and public service key workers. However, our findings should not necessarily be taken to imply that mental health has not been adversely affected amongst health and social care workers. First, this study looked just at symptoms of anxiety and depression and only measured from the start of the first lockdown in 2020. By this point, hospitals were already overwhelmed with COVID-19 patients. As we lack data on mental health in this group prior to the pandemic, it is therefore unclear whether symptoms recorded here were higher than usual baselines. Second, this study looked at general symptoms of anxiety and depression, but other studies have suggested effects on wider aspects of mental health that these measures may not have captured such as post-traumatic stress.^11^ Finally, our category combined the experiences of health and social care workers without differentiating between frontline workers vs those working in other roles that might not have involved exposure to COVID-19 patients. As documented in other studies,^5,6^ the nature of health and social care roles within the pandemic has been shown to have differential effects on mental health. Therefore, future studies are encouraged that explore in more depth the types of health and social care roles that may have been most adversely affected.

It is also notable that key workers in the teacher/childcare sector appeared to have fewer depressive symptoms than non-key workers at the beginning of the pandemic in March 2020 and over the course of follow-up, although the differences were slight. There are several possible explanations for this. First, the start of lockdown in 2020 in the UK involved the closure of schools. This reduced the workload for many in these professions as most schools were unable to deliver a full curriculum online and also reduced changes of exposure to the virus amongst this group. At the same time, teachers did not face the stress of furlough schemes or unemployment as their jobs were recognised as essential when society did reopen, which may contrast with non-key worker sectors where a greater proportion of people were furloughed. Nonetheless, some other workers within this key worker category such as nursery workers did experience furlough and redundancy. As the pandemic continued and schools reopened, teaching and childcare then offered an opportunity to maintain face-to-face social interactions with a population less likely to be physically affected by the virus (children and adolescents),^24^ which could have helped improve their wellbeing during lockdowns and when the strictest physical distancing measures were in place.

Our study did not find any statistical differences between the mental health of public service workers and non-key workers. Many in the former group may enjoy greater job security and lower furlough rates, as well as the ability to work from home. Whilst many non-key workers may have also been able to work from home, they may have been faced with fears of job losses, financial concerns, and stress due to not being able to leave home for work. Although the aim of this study was to report trajectories rather than prevalence, non-key workers evidenced anxiety symptoms which were higher than averages in other studies pre-pandemic using the same measure (2.7-3.2^25^). These findings point to the importance for monitoring and supporting mental health in the population as a whole in the current and in future pandemics.

This study has a number of strengths. It utilised a large sample with sufficient heterogeneity to include good stratification across all major socio-demographic groups. The analyses were weighted on the basis of population estimates of core demographics, with the weighted data showing good alignment with national statistics from the Labour Force Survey;^21^ a nationally representative study. Due to the richness of the dataset, we were able to employ advanced statistical analyses to examine the trajectories of depressive symptoms and anxiety amongst key workers in various sectors since the first lockdown in the UK across different stages of the pandemic over 12 months. Despite these strengths, the limitations of our study raise important points for future search on mental health amongst key workers. First, despite the effort to make our sample representative to the working population in England, there is still the possibility of potential biases due to omitting other demographic factors that could be associated with survey participation in the weighting process. Second, we were only able to analyse data with respondents who reported themselves in government identified ‘key worker’ roles at the start of the pandemic, but the definition of this changed throughout the pandemic. Future study is required to capture how changes in key worker status designation may have impacted mental health of these groups. Moreover, we lacked data on participants’ mental health prior to the COVID-19 pandemic. It therefore remains unclear whether the levels of depressive and anxiety symptoms had already been consistently high amongst the key workers in sectors such as utility, food-chain, transport, and delivery prior to COVID-19, or whether the conditions exacerbated their mental health during the pandemic. Future research is encouraged to look at the longer-term mental health trajectories of key worker in these groups, including after the current pandemic is under control. Although due to our large sample size we were able to examine more key worker categories than prior studies, there was still some heterogeneity within the key worker groups in our study. Due to data limitations, we also were unable to include information on the frontline nature of key workers’ roles, which will be an important confounder to explore in future studies. Finally, we were unable to account for whether and to what extent individual key workers within each category interfaced directly with the public, which might affect their levels of depression and anxiety.^5,6,17^

Our findings indicate that the mental health of individuals in key worker roles such as essential services that have been less visible than health and social care has been worst affected during the current pandemic, suggesting the potential importance of mental health screening in at-risk occupations during pandemics. More mental health support will therefore be needed to deal with these symptoms as the pandemic eases. There is also a need for more fine-grained analyses of the mental health of different types of key workers, particularly those working in utility, transport, and food chain roles to identify individuals in specific occupations who will need this support most. Future research should also seek to understand ways in which workplace measures to mitigate risk may have been inadequate during the current pandemic, to inform policies for future pandemics.

## Data Availability

Anonymous data will be made available in early 2022.

## Declarations

### Ethics approval and consent to participate

Ethical approval for the COVID-19 Social Study was granted by the UCL Ethics Committee. All participants provided fully informed consent. The study is GDPR compliant.

### PPI

The research questions in the UCL COVID-19 Social Study built on patient and public involvement as part of the UKRI MARCH Mental Health Research Network, which focuses on social, cultural and community engagement and mental health. This highlighted priority research questions and measures for this study. Patients and the public were additionally involved in the recruitment of participants to the study and are actively involved in plans for the dissemination of findings from the study

### Author contributions

FB designed the study. FB analysed the data. EP, FB, and KM wrote the first draft. All authors provided critical revisions. All authors read and approved the submitted manuscript.

### Data availability

Anonymous data will be made available in early 2022.

